# Implementation of a Web-Based Symptom Checker to Manage the Quarantine of the USS Theodore Roosevelt Crew Following a Shipboard Outbreak of SARS-CoV-2

**DOI:** 10.1101/2021.08.25.21254738

**Authors:** Robert L. Fenequito, Daniel J. Houskamp, Vincent Siu, Jamal T. Rorie, Nikunj H. Bhatt, Brett Luartes, Maya Kuang, Donald Shell, John Reeder, Katie Rainey, Niels H. Olson

## Abstract

**Introduction:** In late March 2020, the USS Theodore Roosevelt (TR), a nuclear-powered aircraft carrier, pulled into port in the US territory of Guam to assess the severity of a developing outbreak of COVID-19 aboard the ship. A small staff contingent of 60 personnel from US Naval Hospital (USNH) Guam was tasked with the medical care of 4,079 sailors who were placed in single room quarantine amongst 11 hotels across the island of Guam. With the assistance of the Defense Digital Service, the USNH Guam staff implemented a web-based symptom checker, which allowed for monitoring of developing COVID symptoms, and selective testing of symptomatic individuals.

**Materials and Methods:** Sailors from the TR were placed in quarantine or isolation cohorts upon debarking the ship. Sailors not positive for COVID-19 were quarantined amongst 11 hotels on Guam. Sailors positive for COVID-19 were isolated aboard Naval Base Guam (NBG). A retrospective cohort analysis and subgroup analyses were performed on symptom data obtained from sailors in quarantine. The sailors recorded their symptoms and temperature in a web-based symptom checker that assigned a symptom severity score (SSS). Sailors with a SSS >50 were evaluated by a medical provider and re-tested. Data were collected from 4 April 2020 to 1 May 2020. Sailors required two negative tests to exit quarantine and re-embark the ship. The time course, and most common cluster of symptoms associated with a positive COVID-19 PCR test were determined retrospectively after data collection.

**Results:** The web-based symptom checker was successful in establishing daily positive contact and symptom monitoring of susceptible individuals in quarantine. 4,079 sailors in quarantine maintained positive contact with medical staff via the symptom checker, with at least 81% of the sailors recording their symptoms on a daily basis. Individuals with high symptom scores were quickly identified and underwent further evaluation and repeat COVID-19 testing. A cohort of 331 sailors tested positive for COVID-19 while in quarantine and recorded symptoms in the symptom checker before and after a positive COVID-19 test. In this cohort, the most frequent symptoms reported prior to a positive test were headache, anosmia, followed by cough. The symptom of anosmia was reported more frequently in sailors positive for COVID-19, compared to a cohort of matched controls. A small medical staff was able to monitor developing symptoms in a large quarantined population, while efficiently allocating resources, preserving personal protective equipment (PPE), and maintaining isolation and social distancing protocols.

**Conclusions and Relevance:** The application provided a tool for broad health surveillance over a large population while maintaining strict quarantine and social distancing protocols. Highly symptomatic sailors were quickly identified, triaged, and transferred to a higher level of care if indicated. The symptom checker and predictive model generated from the data can be utilized by military and civilian public health officials to triage large populations and make rapid decisions on isolation measures, resource allocation, selective testing.

## Introduction

On 31 December 2020, initial cases of a pneumonia of unknown etiology were first reported to the WHO from Wuhan City, Hubei Province of China.^1^ This illness was found to be caused by a novel coronavirus (SARS-CoV-2), and the disease, termed COVID-19, rapidly spread across the globe and was declared to be pandemic by the WHO on 11 March 2020.^2-3^ The first case aboard the USS Theodore Roosevelt (TR), a San Diego based nuclear-powered aircraft carrier, was reported on 22 March 2020.^4^ Seven more cases were discovered in the following days, and on 27 March 2020, the TR returned to port in Guam to assess the severity of the outbreak.^4-5^

Charged with the triage and medical care of a large quarantine population in 11 hotels, the USNH Guam staff collaborated with the Defense Digital Service (DDS) to develop and implement a progressive web-based application, Coronavirus (COVID-19) Symptom Checker, that allowed real-time reporting and analysis of the onset, progression, and severity of symptoms of the cohort in hotel quarantine. This tool allowed the TR sailors to self-report their symptoms and then assigned a SSS on a twice-daily basis. This information was used by the medical personnel from USNH Guam assigned to the hotels (Task Force Hotel, TFH) to identify sailors who needed further evaluation, medical treatment, and repeat COVID-19 testing.

Based on this data, we performed a retrospective cohort analysis that attempted to define the time course and most common symptoms associated with a positive COVID-19 PCR test. Many studies have retrospectively identified the clinical characteristics and symptomatology preceding a diagnosis of COVID-19. ^6-8^ In the UK, US, and internationally there has been an outpouring of mobile phone and web-based applications that range in function from symptom self-assessment to collection of population-level epidemiologic data. ^9-12^ The symptom checker can be rapidly implemented across various military, civilian and public health settings to generate predictive and actionable health surveillance data. It allows military commanders and public health officials to identify developing symptomatology, COVID-19 and non-COVID-19 related, and make rapid decisions on appropriate measures for quarantine, health surveillance, resource allocation, and testing.

## Methods

### Study Oversight

The study was supported by the Department of Defense (DoD) and designed by the investigators. Sailors were provided a Privacy Act Statement prior to each use of the symptom checker. The data was collected primarily under DoD’s requirement for commanders to be notified of the health status of the members in their command.^13^

### Data Sources

The TR began COVID-19 testing using a Research Use Only assay with an incidental shipboard laboratory capability while the ship was underway in late March 2020. Once the ship ported in Guam, large scale testing to disembark and divide the crew into quarantine and isolation cohorts was started. The laboratory testing was transitioned over a period of days from the shipboard lab, to a land-based system capable of accommodating the projected testing requirement. The result of this testing determined the movement of sailors off the ship. Initially, only sailors who tested positive were debarked from the ship and placed in isolation on NBG. In the following days, this was transitioned to maximum debarkation of all sailors, with positive cases to isolation on NBG, and negative cases to single occupancy quarantine amongst multiple hotels on Guam.

TFH was assigned to the medical care of the hotel quarantine population. The Marine 3^rd^ Medical Battalion, designated Task Force Medical (TFM), deployed from Okinawa to assist with the outbreak and was tasked to oversee the medical care of the positive COVID-19 sailors in isolation on NBG.

The first hotel was loaded out starting on 1 April 2020, and TFH was directed to develop a plan for triaging, evaluating, and treating quarantined sailors. Over the next two weeks, 4,079 sailors were moved to quarantine in ten additional hotels. On 2 April 2020, the Director of DDS contacted one of the authors (NHO) and offered support to USNH Guam in managing the TR outbreak. DDS had an ongoing project, mysymptoms.mil, a symptom checker tool specifically built for COVID-19, and it was proposed that this tool could assist USNH Guam in managing the quarantine population.^14^

The symptom checker was modified for this purpose and a version was deployed on 4 April 2020. The symptom checker was immediately utilized by TFH, in a phased deployment model across eleven hotels, with an eventual total user population of 4,079 quarantined sailors. The quarantined sailors self-reported their symptoms in the application twice daily, and a temperature screening was performed once daily by TFH. Temperature data for sailors in quarantine was logged into the application starting on day four after implementation. The sailors in isolation on NBG did not begin using the application until eight days after initial implementation.

The sailors were able to select from a list of symptoms and conditions weighted based on severity and then summed to create a SSS. DDS and the Office of Assistant Secretary of Defense, Health Affairs (OASD HA) developed the initial scoring for the symptoms and conditions, and these were subsequently changed in the application to reflect ensuing updated CDC guidelines. Table 1 lists the symptoms, conditions, corresponding weights, and likelihood scoring for each version of the application.

**Table 1:** Symptoms, conditions, weights, and likelihood scoring for each version of the application

| Version 1 (4/3/2020) | v1.1 diff (4/7/2020) | v1.2 diff (4/8/2020) | v1.3 diff (4/28/2020) | v1.4 diff (5/1/2020) |
| --- | --- | --- | --- | --- |
| Initial symptom weights | Symptom weights changes | Symptom weights changes | Symptom weights changes | Symptom weights changes |
| Fever +20 | No change |  | No change | Fever +10 |
| Cough +20 |  |  |  |  |
| Shortness of breath +20 |  |  |  |  |
| Chest pain +20 |  |  |  | remove: Chest pain |
| Diarrhea +20 |  |  |  | Diarrhea +5 |
| Body aches +15 |  | Body aches +20 |  | Body aches +10 |
| Headache +10 |  | Headache +20 |  |  |
| Sore throat +10 |  | Sore throat +20 |  |  |
| Fatigue +15 |  |  |  | Fatigue +5 |
| Itchy, red or pink eyes +10 |  |  |  | Itchy, red or pink eyes +5 |
| Altered smell/taste +5 |  |  |  | Altered smell/taste +10 |
| Vomiting +20 |  |  |  | Vomiting +5 |
| Dizziness or lightheadedness +10 |  |  |  | Dizziness or lightheadedness +5 |
|  |  |  | Chills or repeated shaking with chills +5 | Chills or repeated shaking with chills +10 |
| Initial condition weights | Condition weight changes | Condition weight changes | Condition weight changes | Condition weight changes |
| Confusion or trouble staying awake +20 |  |  |  | New confusion or inability to arouse +40 |
| Difficulty breathing +20 | No change | No change |  | Trouble or difficulty breathing +40 |
| Worsening fever or cough +20 |  |  | Added: | Persistent pain or pressure in the chest +40 |
|  |  |  | Bluish lips or face +20 | Bluish lips or face +40 |
|  |  |  | Inability to arouse +20 | Remove:<br>Inability to arouse +20 |
| Likelihood Scoring | Likelihood Scoring | Likelihood Scoring | Likelihood Scoring | Likelihood Scoring |
| Very Low: 0-9 |  | No change | No change | No change |
| Low: 10-29 | Low: 10-19 |  |  |  |
| Medium: 30-49 | Medium: 20-39 |  |  |  |
| High: 50-79 | High: 40-69 |  |  |  |
| Very High: 80+ | Very High: 70+ |  |  |  |

The symptoms and conditions were summed to create a SSS, and then graded to determine the likelihood of a positive COVID-19 diagnosis. TFH performed a follow-on history, physical exam, and obtained vital signs on sailors who had a SSS of greater than 50, or reported symptoms of chest pain, shortness of breath or conditions of persistent pain or pressure in the chest or trouble or difficulty breathing. The sailors in this category were retested for SARS-CoV-2 by PCR from nasopharyngeal swab. If positive, the sailor was moved to isolation quarters on NBG, under the control of TFM for further treatment and observation. Before re-embarking the ship, all TR sailors were required to have a 14-day asymptomatic period and two negative tests.

## Results

### Population

Between 4 April 2020 and 1 May 2020, 4,179 TR sailors in both isolation and quarantine used the application to record their symptoms twice daily. PCR results were available for 4,171 sailors who used the symptom checker; 796 (19.1%) users logged having at least one symptom or condition. There were 115,172 entries logged in the application over the 28-day period. 4,988 entries (4.3%) reported a positive SSS (>0). 10,652 COVID-19 PCR tests were obtained over the 28-day period, with 1,642 (15.4%) returning as a positive result.

Due to lack of standardization with early testing, some tests were identified only with the last initial and last four of the sailor’s SSN or DoD ID, and as a result, could not be traced back to an individual for the purpose of this analysis. 4,079 sailors who were in quarantine used the symptom checker. For the purpose of this analysis, personnel who were not registered in the ship’s official roster were excluded, reducing the quarantine population to 3,773. Regardless of the usage of the symptom checker, 1,113 sailors across quarantine and isolation tested positive. Of these sailors, 911 used the symptom checker. The symptom type and frequency aligned relative to the date of a positive test for these 911 sailors are presented in Figure 1.

**Figure 1:**
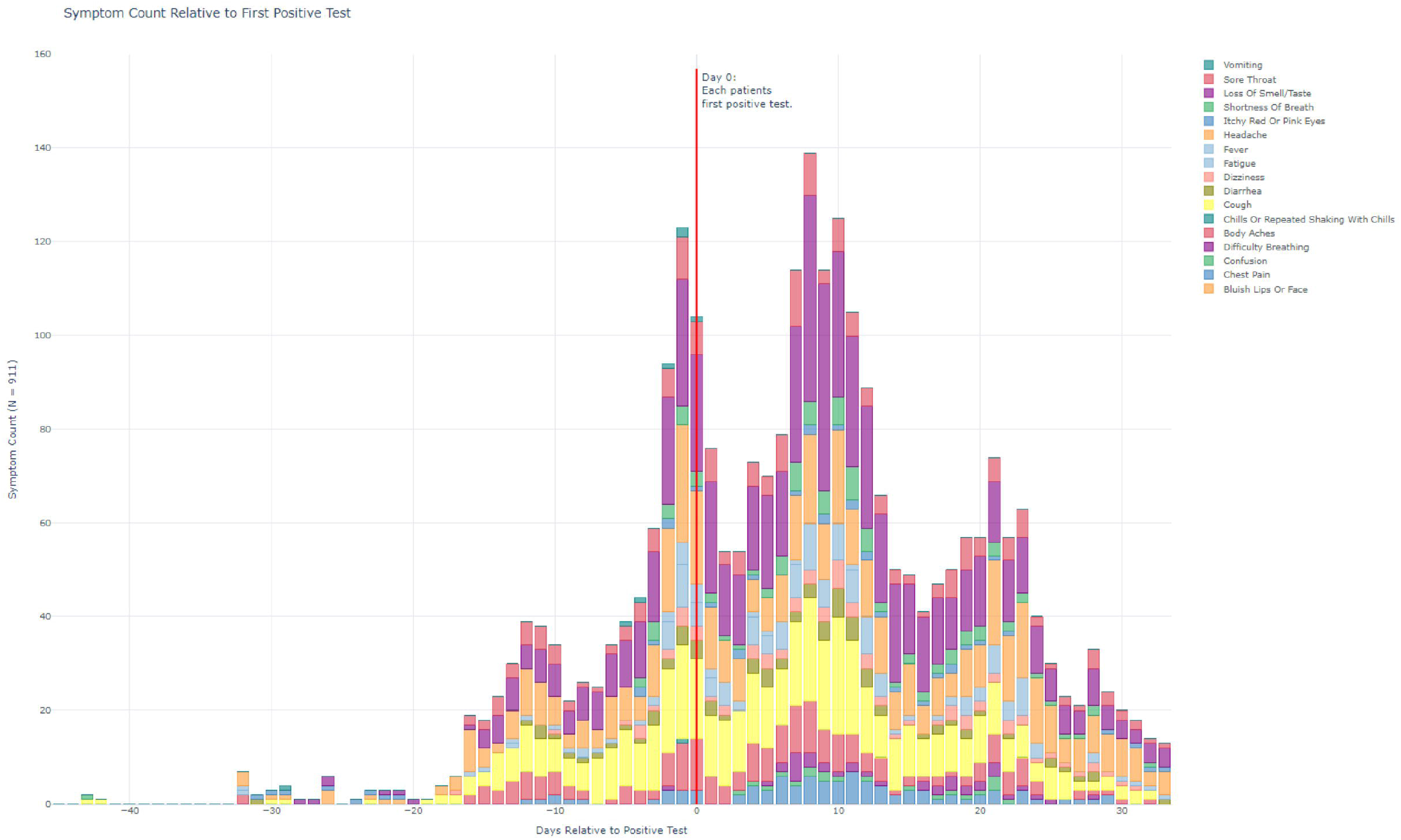
Symptom Type and Frequency Relative to First Positive Test for Cohort of 911 Positive Sailors

The most frequent symptoms reported prior to a positive test in the cohort of 911 sailors were headache, then anosmia, followed by cough. Headache and cough were the two most frequently reported symptoms by sailors who had only negative tests, while fewer than 1% of sailors who had only negative tests reported anosmia. Sailors reported symptoms beyond day 14 of a positive test, and often beyond 30 days after a positive test. Once positive sailors were moved out of hotel quarantine, delay in continued reporting, and loss to follow-up affected the accurate recording of the type and frequency of symptoms. This was reflected in dips in symptom reporting on days 1-6 and 12-20 after a positive test, as seen in Figure 1.

The demographics of the 3,773 sailors in quarantine who used the symptom checker are represented in Table 2. Across this cohort, 537 sailors tested positive. Of those sailors, 331 recorded symptoms in the symptom checker before and after the day of their positive test, in the 28-day window. The most frequent symptoms reported prior to a positive test in this cohort were headache, then anosmia, followed by cough. There was a notable drop-off in symptom reporting in this cohort of sailors by day six after a positive test due to relocation of these sailors to isolation on NBG, and subsequent delay in continued reporting and loss to follow-up. This is reflected in Figure 2. Sailors who were negative for COVID-19 and entered hotel quarantine on the same day as a matched positive case reported the symptom of headache with a similar frequency to positive cases. There was a low frequency of anosmia among the negative matched controls, compared to the positive cohort, as demonstrated in Figure S1 provided in the supplemental materials.

**Table 2:** Demographics of the 4,171 sailors in who reported symptoms and 3781 as quarantine.

|  | Full Population<br>(28 day window) |  | Quarantine Population |  | 331 Positives in Quarantine,<br>and Matched Negatives |  |
| --- | --- | --- | --- | --- | --- | --- |
|  | All | Tested<br>Positive | All | Tested<br>Positive | All | Tested Positive |
| Number | 4179 | 911 | 3773 | 537 | 661 | 331 |
| Sex |  |  |  |  |  |  |
| Male | 3263 (78.23%) | 740(81.23%) | 2913(77.21%) | 409(76.16%) | 514(77.76%) | 256(77.34%) |
| Female | 908 (21.77%) | 171(18.77%) | 860(22.79%) | 128(23.84%) | 147(22.24%) | 75(22.66%) |
| Age (years) |  |  |  |  |  |  |
| Mean (SD) | 27.39 (6.70) | 27.40(6.70) | 27.38(6.69) | 27.44(6.70) | 28.05(6.87) | 27.95(6.91) |
| Range | 18 - 59 | 18 - 49 | 18 - 59 | 18 - 49 | 18 - 52 | 18 - 49 |
| 18-19 | 198(4.75%) | 49(5.38%) | 175(4.64%) | 27(5.03%) | 27(4.08%) | 13(3.93%) |
| 20-29 | 2636(63.20%) | 566(62.13%) | 2389(63.32%) | 332(61.82%) | 396(59.91%) | 203(61.33%) |
| 30-39 | 1090(26.13%) | 241(26.45%) | 987(26.16%) | 147(27.37%) | 190(28.74%) | 91(27.49%) |
| 40-49 | 233(5.59%) | 55(6.04%) | 209(5.54%) | 31(5.77%) | 47(7.11%) | 24(7.25%) |
| 50-59 | 14(0.34%) | 0(0.00%) | 13(0.34%) | 0(0.00%) | 1(0.15%) | 0(0.00%) |
| Max SSS |  |  |  |  |  |  |
| Mean (SD) | 4.52(13.55) | 10.05(20.37) | 3.70(11.43) | 8.21(15.66) | 6.11(13.89) | 8.38(15.54) |
| > 0 | 796(19.08%) | 340(37.32%) | 630(16.70%) | 178(33.15%) | 171(25.87%) | 114(34.44%) |
| > 50 | 65(1.56%) | 37(4.06%) | 40(1.06%) | 13(2.42%) | 11(1.66%) | 7(2.11%) |
| Symptoms |  |  |  |  |  |  |
| Loss Of Smell/Taste | 185(4.44%) | 162(17.78%) | 84(2.23%) | 61(11.36%) | 47(7.11%) | 42(12.69%) |
| Cough | 238(5.71%) | 114(12.51%) | 181(4.80%) | 60(11.17%) | 58(8.77%) | 41(12.39%) |
| Headache | 438(10.50%) | 172(18.88%) | 375(9.94%) | 110(20.48%) | 98(14.83%) | 66(19.94%) |
| Fever | 23(0.55%) | 17(1.87%) | 21(0.56%) | 15(2.79%) | 11(1.66%) | 10(3.02%) |
| Body Aches | 138(3.31%) | 71(7.79%) | 109(2.89%) | 43(8.01%) | 36(5.45%) | 29(8.76%) |
| Diarrhea | 91(2.18%) | 36(3.95%) | 77(2.04%) | 22(4.10%) | 24(3.63%) | 13(3.93%) |
| Sore Throat | 155(3.72%) | 67(7.35%) | 124(3.29%) | 36(6.70%) | 38(5.75%) | 24(7.25%) |
| Dizziness | 76(1.82%) | 32(3.51%) | 61(1.62%) | 18(3.35%) | 14(2.12%) | 9(2.72%) |
| Fatigue | 100(2.40%) | 60(6.59%) | 73(1.93%) | 33(6.15%) | 22(3.33%) | 18(5.44%) |
| Chest Pain | 63(1.51%) | 28(3.07%) | 50(1.33%) | 15(2.79%) | 13(1.97%) | 9(2.72%) |
| Shortness of Breath | 56(1.34%) | 30(3.29%) | 38(1.01%) | 12(2.23%) | 7(1.06%) | 4(1.21%) |
| Difficulty Breathing | 33(0.79%) | 14(1.54%) | 20(0.53%) | 1(0.19%) | 4(0.61%) | 1(0.30%) |
| Itchy Red Or Pink Eyes | 46(1.10%) | 14(1.54%) | 40(1.06%) | 8(1.49%) | 5(0.76%) | 3(0.91%) |
| Vomiting | 19(0.46%) | 5(0.55%) | 19(0.50%) | 5(0.93%) | 6(0.91%) | 3(0.91%) |
| Chills Or Repeated Shaking With Chills | 19(0.46%) | 1(0.11%) | 19(0.50%) | 1(0.19%) | 4(0.61%) | 1(0.30%) |
| Confusion | 11(0.26%) | 5(0.55%) | 6(0.16%) | 1(0.19%) | 0(0.00%) | 0(0.00%) |
| Bluish Lips Or Face | 0(0.00%) | 0(0.00%) | 0(0.00%) | 0(0.00%) | 0(0.00%) | 0(0.00%) |

**Table 3:**
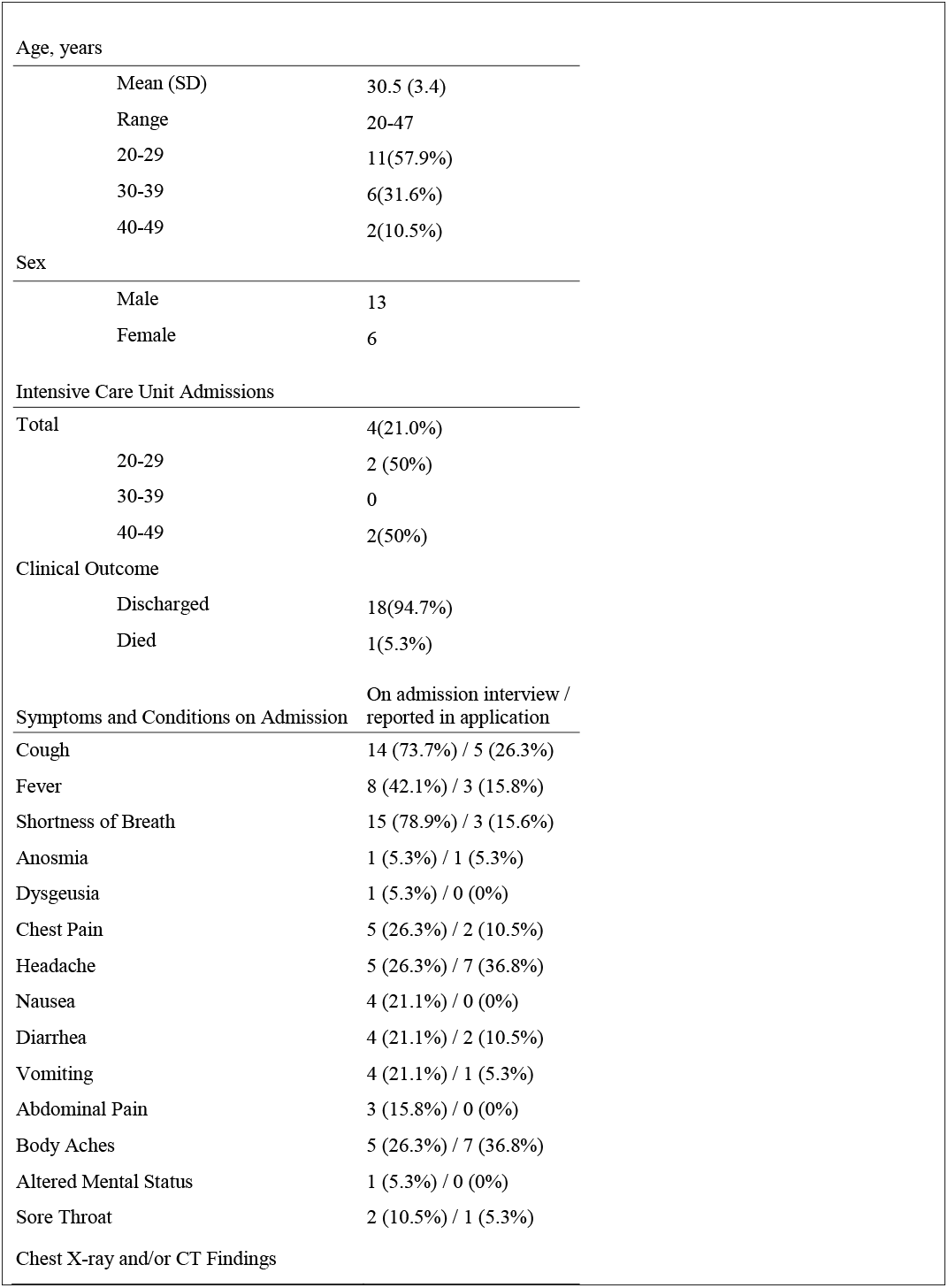
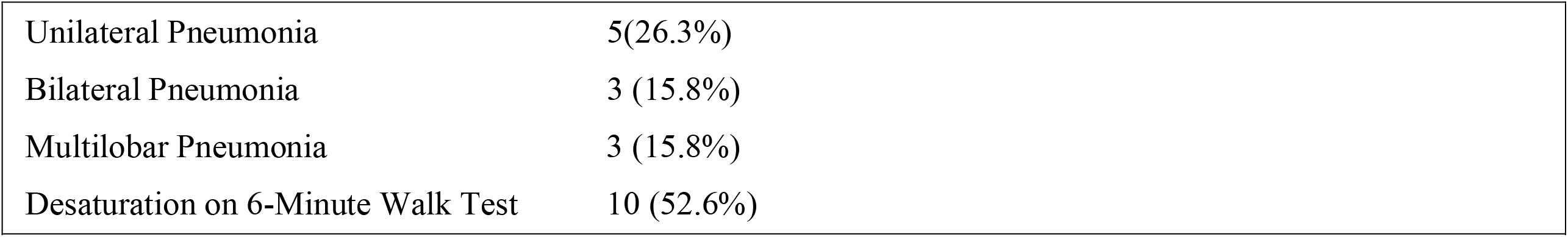
Demographics of sailors admitted to USNH Guam

|  |  |  |
| --- | --- | --- |
| Age, years |  |  |
|  | Mean (SD) | 30.5 (3.4) |
|  | Range | 20-47 |
|  | 20-29 | 11(57.9%) |
|  | 30-39 | 6(31.6%) |
|  | 40-49 | 2(10.5%) |
| Sex |  |  |
|  | Male | 13 |
|  | Female | 6 |
| Intensive Care Unit Admissions |  |  |
| Total |  | 4(21.0%) |
|  | 20-29 | 2 (50%) |
|  | 30-39 | 0 |
|  | 40-49 | 2(50%) |
| Clinical Outcome |  |  |
|  | Discharged | 18(94.7%) |
|  | Died | 1(5.3%) |
| Symptoms and Conditions on Admission | On admission interview /<br>reported in application |  |
| Cough | 14 (73.7%) / 5 (26.3%) |  |
| Fever | 8 (42.1%) / 3 (15.8%) |  |
| Shortness of Breath | 15 (78.9%) / 3 (15.6%) |  |
| Anosmia | 1 (5.3%) / 1 (5.3%) |  |
| Dysgeusia | 1 (5.3%) / 0 (0%) |  |
| Chest Pain | 5 (26.3%) / 2 (10.5%) |  |
| Headache | 5 (26.3%) / 7 (36.8%) |  |
| Nausea | 4 (21.1%) / 0 (0%) |  |
| Diarrhea | 4 (21.1%) / 2 (10.5%) |  |
| Vomiting | 4 (21.1%) / 1 (5.3%) |  |
| Abdominal Pain | 3 (15.8%) / 0 (0%) |  |
| Body Aches | 5 (26.3%) / 7 (36.8%) |  |
| Altered Mental Status | 1 (5.3%) / 0 (0%) |  |
| Sore Throat | 2 (10.5%) / 1 (5.3%) |  |
| Chest X-ray and/or CT Findings |  |  |

|  |  |
| --- | --- |
| Unilateral Pneumonia | 5(26.3%) |
| Bilateral Pneumonia | 3 (15.8%) |
| Multilobar Pneumonia | 3 (15.8%) |
| Desaturation on 6-Minute Walk Test | 10 (52.6%) |
| **Table 3:** Demographics of sailors admitted to USNH Guam |  |

**Table 4.** 4A: Performance metrics for the decision tree selected from a random forest. 4B: relative importance of each parameter in the model.

| **4A** | Sensitivity | Specificity | F1 score | Support |
| --- | --- | --- | --- | --- |
| Sailor will remain negative | 0.71 | 0.79 | 0.75 | 31 |
| Sailor will test positive | 0.77 | 0.69 | 0.73 | 26 |
| Accuracy |  |  | 0.74 | 57 |
| Macro average | 0.74 | 0.74 | 0.74 | 57 |
| Weighted average | 0.74 | 0.74 | 0.74 | 57 |

| **4B** | Relative Importance |
| --- | --- |
| Loss Of Smell/Taste | 0.209 |
| Cough | 0.095 |
| Headache | 0.087 |
| Fever | 0.084 |
| Body Aches | 0.071 |
| Diarrhea | 0.067 |
| Sore Throat | 0.067 |
| Dizziness | 0.064 |
| Fatigue | 0.053 |
| Chest Pain | 0.052 |
| Shortness Of Breath | 0.043 |
| Itchy Red Or Pink Eyes | 0.034 |
| Difficulty Breathing | 0.030 |
| Vomiting | 0.022 |
| Chills Or Repeated Shaking With Chills | 0.019 |
| Confusion | 0.003 |
| Bluish Lips Or Face | 0.000 |

**Figure 2:**
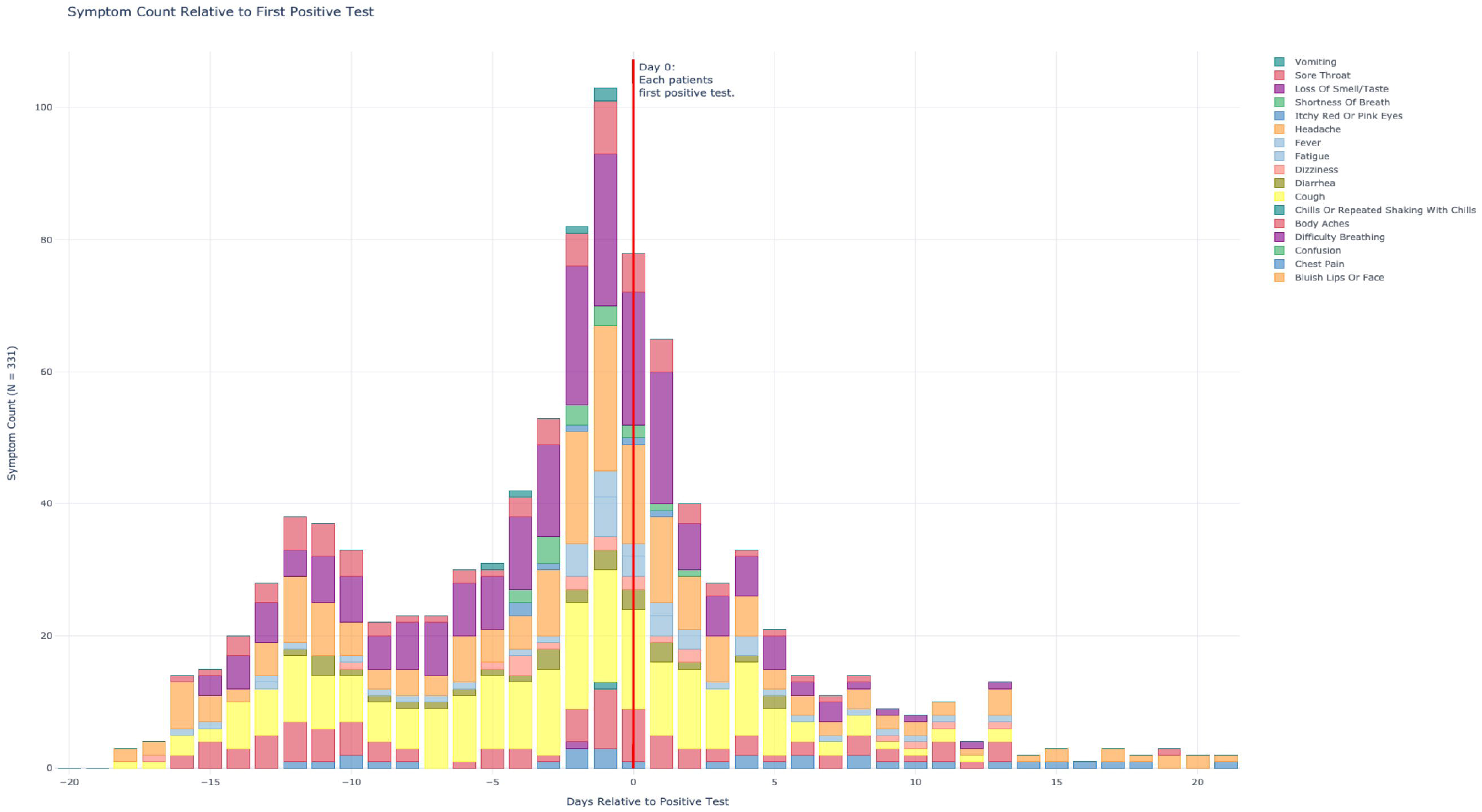
Symptom Type and Frequency Relative to First Positive Test for Cohort of 331 Positive Sailors in Hotel Quarantine

**Figure 3:**
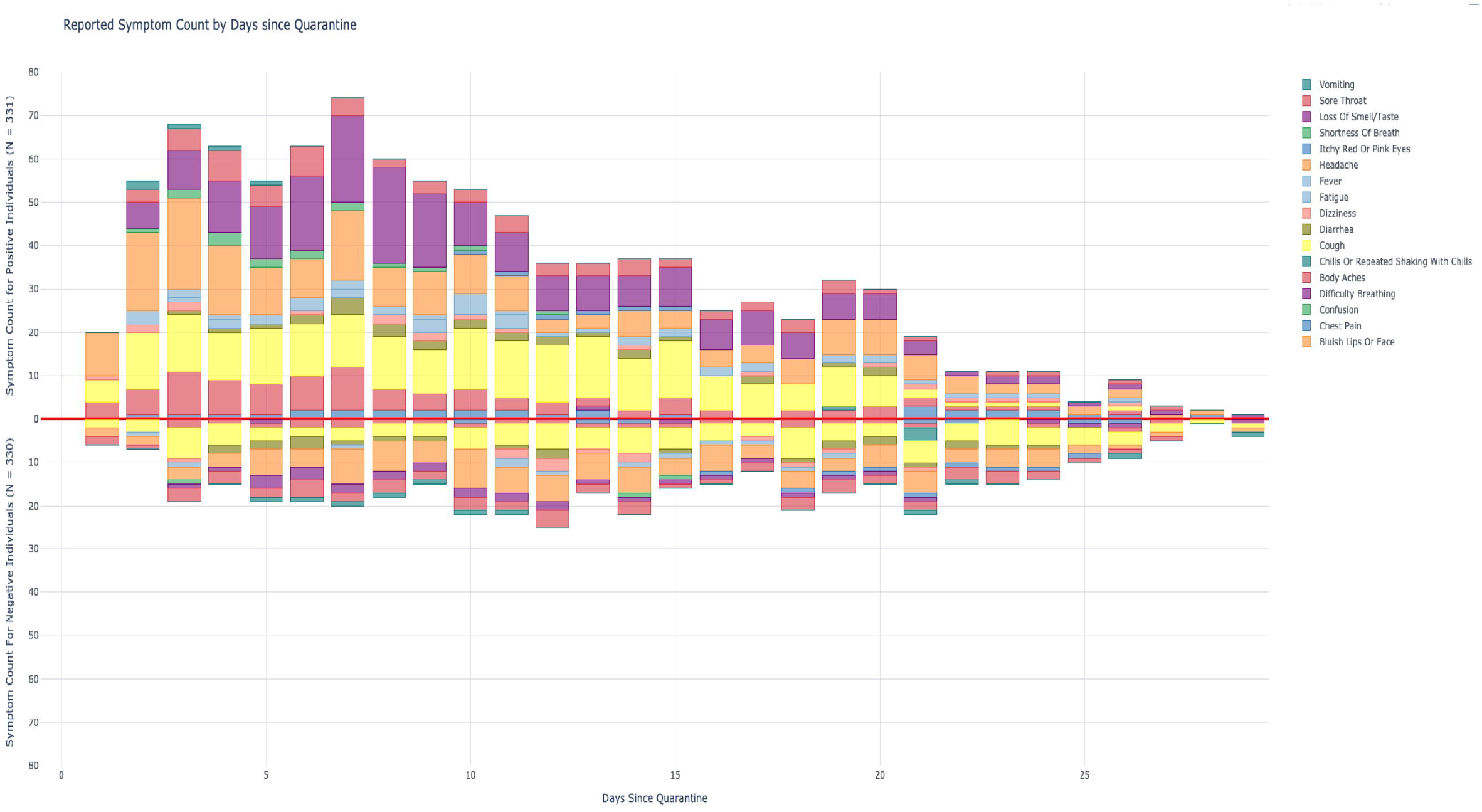
Symptom Type and Frequency of 331 Positive Sailors in Hotel Quarantine, with Negative Controls Matched by Date of Entry into the Same Hotel.

**Figure 4:**
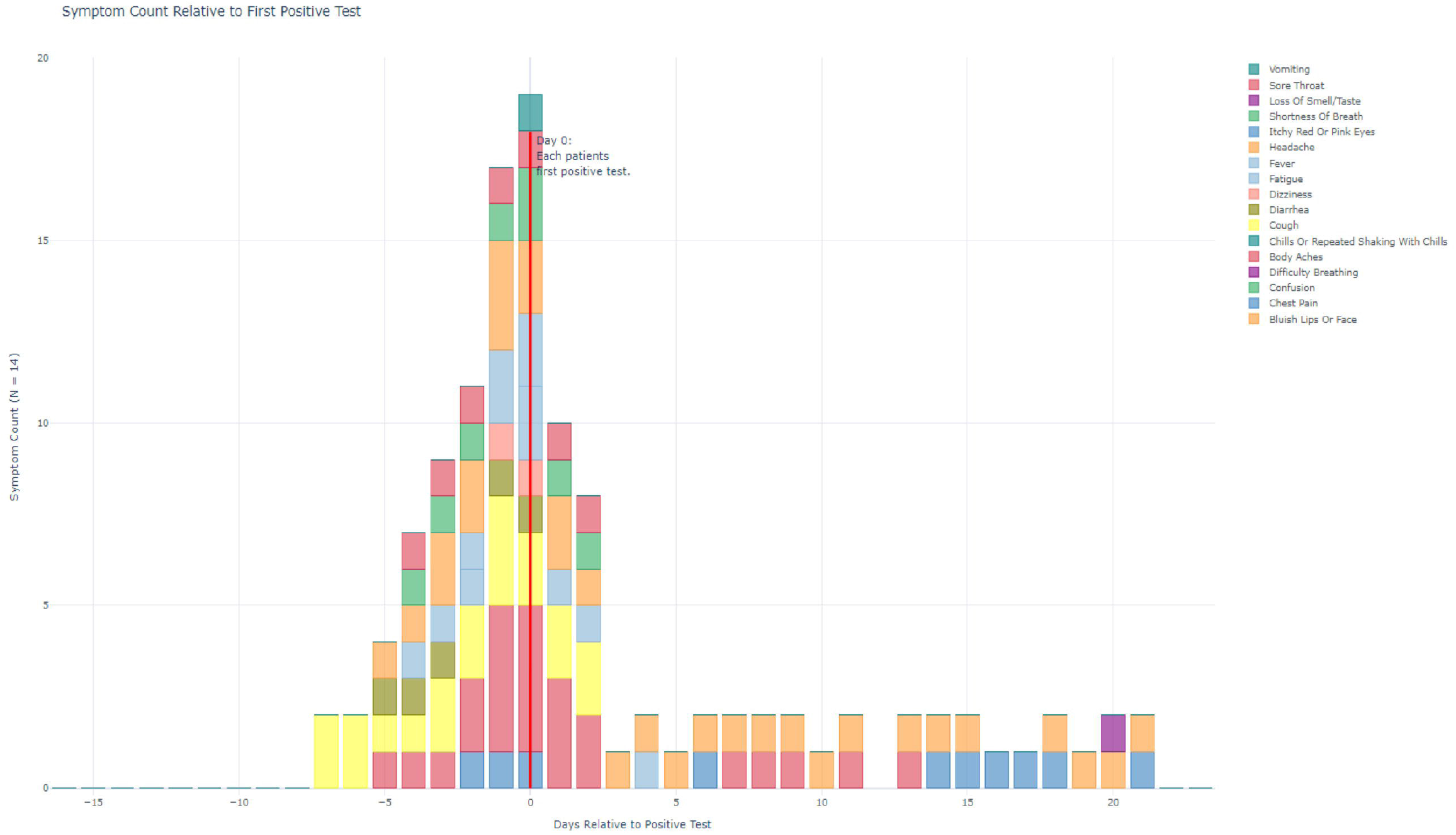
Symptom Type and Frequency of Relative to First Positive Test for Cohort of 14 Positive Sailors that Used the Symptom Checker and were Admitted to USNH Guam

### Inpatient cohort

19 out of 1,113 (1.8%) TR sailors who were positive for COVID-19 from a period of 4 April 2020 and 1 May 2020 required admission to the USNH Guam inpatient wards for further monitoring and observation. A low threshold for inpatient admission was set to capture all patients demonstrating worsening clinical symptomatology and progression of disease. Any sailor who was COVID-19 positive and had abnormal vital signs, findings consolidation on chest x-ray, or abnormal laboratory studies was admitted for observation. Of these 19 sailors, nine were from hotel quarantine, and ten were from isolation on NBG. The age range of sailors was 20 to 47 years, with a mean of 30 years, median 29 years. The average length of admission was 7.5 days (median 5.5), with a minimum of two days and a maximum of 16 days. Four required ICU level care during their admission, and there was one death. For patients aged 30 or over, the average length of hospital stay was 7.4 days (median 6). The demographics of the sailors admitted to USNH Guam is represented in Table S1 of the supplemental materials. The maximum SSS amongst the sailors who required admission was 125. Seven of the admitted sailors had a maximum SSS of 40 or greater. The symptom type and frequency aligned relative to their first positive test for the 14 sailors admitted to USNH Guam who used the symptom checker is reflected in Figure S2 in the supplemental materials.

### Model development

A subset analysis was performed after standardizing all data on v1.4 scoring, for sailors who reported symptoms above a minimum score (SSS > 9) in the window between 4 April 2020 (the first day the symptom checker was deployed to the quarantine population) and 1 May 2020 (the first day of mass re-embarkation). One hotel was excluded from this analysis because it was populated under a different policy. The subset, which contained 141 positive sailors, was matched to 141 sailors who remained negative and reported symptoms above the same minimum score (SSS > 9). The symptom reports were further condensed by collecting each symptom reported by an individual in the specified window into a single entry, thus each example row corresponds to an individual and which symptoms they reported during the window along with their ultimate test result. The set of 282 sailors was split into 80%/20% sets to be used to train and test a random forest classifier to predict whether each sailor tested positive. The results of the unoptimized classifier^15^ are reported in Table S2 of the supplemental materials.

## Discussion

Similar to many other healthcare facilities around the world, in early March 2020, USNH Guam was attempting to develop triage, treatment, logistics, and disposition protocols for the management of large volumes of patients exposed to COVID-19. With the arrival of the USS Theodore Roosevelt on 27 March 2020, USNH Guam was faced with an overwhelming task of managing an outbreak among 4,778 sailors while maintaining normal hospital operations. The development of the symptom checker by the DDS allowed the USNH Guam staff to closely monitor the clinical symptomatology of sailors in single occupancy quarantine, and allowed selective identification and retesting of highly symptomatic individuals.

Among the 911 patients from the TR who were positive for COVID-19 and used the symptom checker, the three most commonly reported symptoms that preceded a positive COVID-19 test were headache, anosmia, and cough. Among 331 sailors who tested positive while in quarantine and recorded symptoms in the checker before and after a positive test, the most frequent symptoms reported prior to a positive test were headache, followed by anosmia and cough. Of the sailors who tested positive in this cohort of 331, 65.6% reported a SSS of 0 in the symptom checker. The asymptomatic rate across all 911 sailors who tested positive was 62.7%, suggesting a higher rate of asymptomatic carriage compared to the general population.^16^ Symptoms in positive sailors persisted beyond 14 days, and in some cases, beyond 30 days.

Comparing the 331 positive cases matched to negative cases that entered quarantine on the same day, the symptom of headache occurred with roughly double the frequency in the matched controls, suggesting that this symptom is somewhat specific for COVID-19. The symptom of anosmia was reported more frequently in sailors positive for COVID-19, suggesting it is a more specific symptom for the disease, similar to findings published by the COPE Consortium.^10-11^ These findings may assist clinicians in focusing in on relevant symptomatology in patients presenting with non-specific flu-like symptoms.

Of the 1,113 sailors who were positive for COVID-19, 19 sailors (2%) required admission, which was less than US hospitalization rates reported by the CDC for a similar age demographic during the same time period.^17^ Patients under the age of 30 tended to have similar length hospital stays to patients over 30 years old, on average, 7.5 days, versus 7.4 days for patients over 30 years old. Patients over the age of 30 had higher rates of ICU admission (25%) than those less than 30 years old (18%). Both patients over the age of 40 required ICU-level care during their hospital stay. In the initial history for patients admitted to the hospital, the three most common reported symptoms on admission interviews were shortness of breath, cough, and fever. Anosmia and dysgeusia were reported in only two patients’ admission histories. The three most common symptoms reported on the symptom checker among the admitted sailors were body aches, headache, and cough. Only one of the admitted sailors reported anosmia on the symptom checker.

The use of the web-based symptom checker allowed monitoring and evaluation of symptom scores in real-time and allowed TFH to quickly allocate limited medical resources to floors of hotels, where sailors required care, repeat COVID-19 testing or, if indicated, transfer to a higher level of care. By selectively choosing to evaluate sailors with high symptom scores, TFH was able to limit their exposure, preserve PPE, maintain social distancing protocols, and maintain a single-occupancy quarantine model in each hotel.

The management of the USS Theodore Roosevelt COVID-19 outbreak demonstrated a successful initial use case of the DDS symptom checker, and the application is currently being deployed in ongoing military health operations. The web application, accessible through mobile devices, can be rapidly implemented. The application was tested, piloted, and fully deployed in four days to an eventual population of 4,179 sailors. The application also allows for the strategic deployment of medical staff, resources, and testing capabilities to allow small medical contingents to deliver efficient care to exponentially larger populations. The symptom checker provides real-time health surveillance of large cohorts, allowing military commanders and, potentially, civilian public health officials to quickly analyze symptom trends, provide focused testing, and identify potential outbreak hotspots to limit further disease spread.

### Limitations

We did identify some limitations in the use of the web-based symptom checker. In the early months of the pandemic, there was limited understanding of COVID-19 regarding typical symptomatology and progression of the disease. As a result, symptoms and score weights were chosen and revised as COVID-19 guidelines were updated. In the setting of the application, the model may be a useful revision to the current expert-designed scoring system; however, clinical assessment may at times differ from the model’s prediction, as the model’s prediction may not align with any particular sailor’s symptoms in a clinical setting. Although the rates of hospitalization amongst the TR sailors (∼2%) are lower than that of the US population, the TR population’s demographics do not reflect the general population. The majority of the TR sailors were young, healthy, 18 to 25-year-olds without comorbidities. Additionally, the progression of symptoms through the full course of the disease was not able to be accurately described, due to the aggregate effect of loss to follow up, delayed onset of reporting, and lack of continued compliance with the application due to poor wireless connectivity once positive sailors were transported to isolation on NBG.

## Conclusion

After completing the remainder of her deployment, the USS Theodore Roosevelt returned to its homeport of San Diego on 9 July 2020, without further SARS-CoV-2 outbreaks.^18^ This initial use case demonstrates that a web-based symptom checker can be rapidly and effectively implemented on a remote, resource limited island, 6,000 miles from the continental US. We believe it can provide military commanders, public health officials, and local and state governments real-time access to predictive and actionable health surveillance data to quickly identify developing symptomatology, COVID-19 and non-COVID-19 related, which will assist in making rapid decisions on quarantine measures, resource allocation, and testing.

## Data Availability

Copyright Statement: I am a military service member or employee of the United States government. This work was prepared as part of my official duties. Title 17 U.S.C. 105 provides that 'copyright protection under this title is not available for any work of the United States Government.' Title 17 U.S.C. 101 defines a U.S. Government work as work prepared by a military service member or employee of the U.S. Government as part of that person's official duties.

## Acknowledgements

The authors wish to thank Captain Maria A. Young, RN, Captain Christopher Jack, RN, Lieutenant Kyu Choi, DDS, the Physicians, Dentists, Nurses, and Corpsmen of Task Force Hotel and the staff of US Naval Hospital Guam; Brett Goldstein, Stuart Casarotto, Patrick Dufour, Marie Thacker, Jeff Hackshaw, Elliott Wilkes, and the staff of Defense Digital Service; Colonel William O’Connell, MD, US Army, Jangwoo Lee, PhD, US Army, and the laboratory staff of Brian Allgood Army Community Hospital; Charles Yetman, Chris Mayfield, and the mathematicians, scientists, and engineers of Naval Information Warfare Command; Elaine E. Thompson of Henry M. Jackson Foundation for Military Medicine; Alan Nordholm, PhD, of Uniformed Services University; Rear Admiral John Menoni, US Navy, and the staff of Joint Region Marianas, Vice Admiral William R. Merz, US Navy, Captain Chris Sears, MD, US Navy, and the staff of the US Seventh Fleet; Admiral John C. Aquilino, Captain Michael McGinnis, MD, US Navy, and the staff of the US Pacific Fleet; Rear Admiral Louis Tripoli, MD, US Navy, US Indo-Pacific Command, Rear Admiral (retired) William Roberts, MD, and the members of the Indo-Pacific Research Alliance for Military Medicine; the people of Guam; and most especially, the crew of the USS Theodore Roosevelt, qui plantavit curabit.

